# Handgrip strength on the unaffected side associated with cardiorespiratory fitness in male patients with stroke

**DOI:** 10.1101/2023.05.23.23290430

**Authors:** Yuan Chen, Mingchao Zhou, Fubin Zha, Shaohua Zhang, Jiao Luo, Meiling Huang, Qiangqing Yang, Linlin Shan, Yulong Wang

**Affiliations:** The First Affiliated Hospital of Shenzhen University, Shenzhen Second People’s Hospital, Shenzhen Institutes of Advanced Technology, Chinese Academy of Sciences, Shenzhen, Guangdong, China; Shenzhen Dapeng New District Nan’ao People’s Hospital, Shenzhen, China

**Keywords:** Cardiorespiratory fitness, handgrip strength, muscle strength, stroke

## Abstract

**Objectives:** To investigate factors related to cardiorespiratory fitness in patients with stroke and explore the association between handgrip strength (HS) and peak oxygen utilization (VO_2_peak).

**METHODS:** The present study adopted a cross-sectional method. Seventy male patients who had been clinically diagnosed with ischemic or hemorrhagic stroke were recruited for this study. HS on the unaffected side (uHS) was measured using a hydraulic hand dynamometer and adjusted for body mass index (uHS_BMI_) and body surface area (uHS_BSA_). Concurrently, the VO_2_peak was measured using a cardiopulmonary exercise test system. Univariate, multiple linear regression analyses were used to evaluate the association between various participant characteristics and the VO_2_peak.

**RESULTS:** The average age of the 70 selected male patients was 51.6 ± 10.3 years. The Barthel Index (BI), uHS, uHS_BMI_, and uHS_BSA_ were the independent predictors of VO_2_peak. The National Institutes of Health Stroke Scale (NIHSS), body mass index (BMI), and body surface area (BSA) were negatively correlated with the VO_2_peak. The estimation of VO_2_peak using linear regression, including age, BI, uHS_BSA_, and anaerobic threshold (AT) as independent variables, explained 65.5% of the variance in the VO_2_peak.

**CONCLUSION:** BMI- and BSA-adjusted uHS appear to be independent factors associated with cardiorespiratory fitness in male patients with stroke. The anaerobic threshold (AT) combined with uHS_BMI_/uHS_BSA_ may provide a more reliable assessment of the aerobic capacity post-stroke. The measurement of handgrip strength is a simple, risk-stratifying method that may help determine the cardiorespiratory fitness of patients with stroke, but a larger study with diverse subjects is needed.

## INTRODUCTION

Stroke is the second most common cause of death and the third major cause of death and disability worldwide^[1]^. Many stroke survivors suffer residual impairments, such as hemiparesis, aphasia, dysarthria, and cognitive dysfunction, which impair the ambulatory and activity of daily living (ADL) functions^[2–5]^. Compared to age- and sex-matched healthy individuals, patients with stroke have significantly lower cardiorespiratory fitness (CRF)^[6]^. Patients with lower CRF levels may tend to avoid physical activity, which further impairs their ability to perform daily activities and maintain independent functioning.^[7]^ Reduced CRF can also increase the risk of recurrent cerebrovascular events. Therefore, it is crucial to understand the factors related to CRF in patients with stroke to develop appropriate therapies that can improve their physical activity levels and prevent the further reduction of function.

Previous studies have investigated some factors associated with post-stroke CRF. Peak oxygen uptake (VO_2_peak) is the most common measure used to assess aerobic capacity in stroke research.^[6]^ Age and BMI were found to be negatively associated with the VO_2_peak after stroke.^[8]^ A significant positive relationship was observed between VO_2_peak and the activity of daily living (ADL) function, measured using the Barthel Index.^[7]^ Information about the influencing factors on CRF may help identify individuals at risk of increased recurrent cerebrovascular events and reduced aerobic capacity. However, the combined effects of these influencing factors on CRF remain unclear. Moreover, some influencing factors are difficult to measure directly. The present study aimed to explore simple influencing factors on CRF in patients with stroke without using time-consuming and cost-intense methods. The goal was to help easily identify individuals at a higher risk of deconditioning and develop appropriate therapies.

CRF and muscle strength are strong risk factors for mortality, and their combination was associated with an increased risk of stroke, suggesting a potential correlation between CRF and muscle strength.^[9,10]^ Handgrip strength (HS), as a simple, easy, and inexpensive way to evaluate muscle strength, is an emerging prognostic index associated with adverse cardiovascular outcomes.^[11]^ Previous studies on patients with paraplegia, chronic obstructive pulmonary disease, congestive heart failure, and acquired immune deficiency syndrome have reported a significant correlation between HS and VO_2_peak (p < 0.05).^[8,12–15]^ Meanwhile, our previous study has shown that body mass index (BMI)- and body surface area (BSA)-adjusted HS can be used to identify the normal VO_2_max in healthy young adults, particularly in males.^[8]^ However, the correlation in patients with stroke is unknown. Here, our research objective was to investigate the association between HS-related factors and cardiorespiratory fitness (VO_2_peak) in patients with stroke. Muscle strength was hypothesized to contribute to poorer fitness.

## Methods

### Study Design

This study used a cross-sectional and prospective design. The study protocol was approved by the research ethics committee of Shenzhen Second People’s Hospital (approval number: 20200601045-FS01) and registered at the China Clinical Trial Center (ChiCTR2000035308). The study was conducted in accordance with the Declaration of Helsinki. Data were fully anonymized prior to analysis.

### Participants

In this study, we focused exclusively on males, because our previous study showed that the association between HS_BMI_/HS_BSA_ and CRF was strong in young male adults but weak in young female adults. To achieve 80% power and detect an effect size (Cohen’s f^2^) of 0.2 attributable to 4 independent variables using an F Test (multiple regression analysis) with a significance level (alpha) of 0.05, a minimum sample size of 65 participants was determined using the statistical software package G*power. Seventy-three male patients with stroke participated in this cross-sectional study from the rehabilitation center of Shenzhen Second People’s Hospital, Shenzhen City, China, between May 2020 and November 2021. Of these, three participants had missing CPX parameters, because they could not exercise to a sufficient intensity to attain the peak VO_2_. The final cohort comprised 70 men for whom complete information, including AT, VO_2_peaks, and the primary outcomes were available. The inclusion criteria were (1) diagnosis of ischemic or hemorrhagic stroke confirmed by CT or MRI, (2) age 18 years or greater, (3) 48 h post-stroke, (4) single-hemisphere stroke, (5) presence of some upper-extremity voluntary activity, as indicated by the ability to move the proximal and/or distal joints against gravity; (6) ability to comprehend simple test instructions. The exclusion criteria were (1) other neurological disorders that resulted in permanent damage, (2) inability to perform a maximal exercise test in accordance with the absolute contraindications for maximal exercise testing by the American College of Sports Medicine(ACSM), and (3) absence of informed consent. Before data collection, all participants were informed of the objectives and methodology of the study and provided a written informed consent form.

### Demographic and Anthropometric Factors

Demographic characteristics, such as sex and age, were recorded after admission. Anthropometric factors such as height (m), weight (kgr), body mass index (BMI, kg/m^2^), and body surface area (BSA) were determined. BMI was calculated as body weight/height in meters squared (kg/m^2^). BSA was calculated using the Mosteller formula: 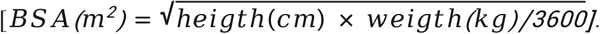.

### Clinical History and Stroke-Related Factors

Physician-diagnosed clinical diseases, including hypertension and diabetes mellitus (DM), were self-reported by each participant. Hypertension was confirmed if the participant had any or all of the following conditions: physician-diagnosed hypertension, mean systolic blood pressure (BP) ≥ 140 mmHg, mean diastolic BP ≥ 90 mmHg, prescription of anti-hypertensive drugs^[16]^. According to the international standard, DM was defined by any or all of the following conditions: fasting plasma glucose ≥ 7.0 mmol/L, random plasma glucose ≥ 11.1 mmol/L, glycated hemoglobin (HbA1c) ≥ 6.5%, self-reported DM diagnosed by a physician, prescription of glucose-lowering drugs or insulin treatment^[17]^. Stroke severity on admission to the rehabilitation center was assessed using the National Institutes of Health Stroke Scale (NIHSS) and Barthel index (BI). The type of stroke (ischemic or hemorrhagic) and stroke location (left or right) were also recorded.

### Grip Strength Measurement

According to standard procedures, the HS of the patients on the unaffected sides was measured using a hydrostatic hand dynamometer (Jamar, 1516801, Patterson Medical, UK)^[18]^. The HS was measured three times, and the mean value was used for the analysis. Each trial was separated by a rest period of at least 30 s. Participants were seated with the shoulder adducted and neutrally rotated, the elbow flexed to 90°, and the forearm and wrist in a neutral position. Absolute HS was defined as an average of the three measurements on the unaffected side. Some studies showed that understanding the association between HS and cardiovascular health is limited by the confounding body size and body composition^[19]^. Therefore, relative HS (HS adjusted for BMI or BSA) has been used to reduce the influence of heterogeneity on the results. Relative HS was reported as the absolute HS divided by the BMI and BSA, uHS_BMI,_ and uHS_BSA_, respectively.

### Cardiopulmonary Exercise Test

CRF was measured in a symptom-limited graded cycle ergometer test on an electronically braked, upright stationary bicycle (MasterScreen; Ergoline, Germany) under the supervision of a physician. Cardiovascular stability was monitored continuously using a 12-lead ECG. The gases were calibrated with standard gases before each test. Expired air was collected breath-by-breath and analyzed using the Jaeger Carefusion system (V-706575; Jaeger, Germany). After 3 min of familiarization, the participants began cycling at a work-load of 20 W, with stepwise work-load increments of 10 W/min, until maximal effort was achieved. Participants pedaled at 60 revolutions per min (rev·min^-1^). Tests were terminated when one or more of the following were observed: (a) respiratory exchange ratio (RER) ≥ 1.1; (b) peak heart rate within ± 10 beat·min^-1^ of the age-predicted maximal value; (c) abnormal blood pressure response or ECG reading; or (d) inability to continue pedaling above 50 rpm.

The oxygen consumption (VO_2_), carbon dioxide production (VCO_2_), minute ventilation (VE), and respiratory exchange ratio (RER) were determined. Peak oxygen consumption (VO_2_peak) was defined as a 20-60 s averaged value of the last completed increment. The anaerobic threshold represents the submaximal VO_2_, in which a nonlinear rise in VE and VCO_2_ is observed. The oxygen uptake efficiency slope (OUES) is a metric that expresses the ventilatory requirement at a given VO_2_, derived from the relationship between VO_2_ (plotted on the y-axis) and the log transformation of VE (x-axis). The VE versus VCO_2_ slope was calculated by linear regression, excluding the nonlinear part of the data after the onset of ventilatory compensation for metabolic acidosis. Other descriptive measures of exercise testing were peak heart rate, maximal workload, respiratory exchange ratio peak, blood pressure at maximum work-load, and metabolic equivalent at maximum work-load.

### Statistical Analyses

Continuous variables with a normal distribution are expressed as mean ± standard deviation (SD). Categorical variables are expressed in terms of frequency or percentage. The sample size was calculated based on the recorded numbers and reference to data from an earlier study. Participants with missing important data (e.g., uHS, VO_2_peak) were excluded from the final analysis. Secondary indicators that were partially missing were replaced with a mean value. The correlation between VO_2_peak and other characteristics was analyzed by Pearson or Spearman analysis. To investigate independent predictors of VO_2_peak, relevant variables were included in age-adjusted linear regression models. Multivariate analysis was performed to generate a predictive model for VO_2_peak. Variables (clinical characteristics, uHS_BMI_, uHS_BSA_, and AT) were entered into the model. To avoid potential multicollinearity, once a variable had been as an adjusting factor for other variables, it was not included as a covariate in the multivariate linear regression analysis. The intraclass correlation coefficient was calculated to assess the reliability of the calculated VO_2_peak using the regression model. The difference between the measured and calculated VO_2_peak was also evaluated using a Student’s paired *t*-test. In addition, we assessed the agreement between the measured and calculated VO_2_peak using Bland-Altman analysis (measurement difference plotted against mean values within the 95% limits of agreement quantified as the mean difference ± 1.96 × standard deviation difference) and the absolute percent error calculated for each subject as the absolute value of ([actual VO_2_peak – estimated VO_2_peak]/actual VO_2_peak) × 100%^[20]^. Analyses were performed and figures were acquired using SPSS version 21.0 (IBM Corp., Armonk, NY, USA). Two-sided *p* values < 0.05 were considered statistically significant.

## RESULTS

### 3.1. Characteristics of Selected Participants

Seventy patients with stroke participated in this study. Their descriptive, clinical, and anthropometric characteristics are presented in Table 1. Based on the type of stroke, 29 (40.85%) patients experienced hemorrhagic stroke and 42 (59.15%) patients experienced ischemic stroke. Thirty-seven patients (52.11%) had a left lesion, whereas 34 patients (47.89%) had a right lesion. The mean (SD) NIHSS at admission to the rehabilitation unit was 3.32 ± 2.82. Fifty-three participants (74.65%) and 24 participants (33.80%) had hypertension and diabetes mellitus, respectively, as comorbidities. All males were overweight (BMI > 24). Results from the treadmill test showed a mean VO_2_peak ± SD of 16.12 ± 3.69 mL/kg·min. The average value of uHS was 29.80 ± 9.28 kg. In addition, the mean score of BI indicated that the patients had moderate dependence (BI 70.77 ± 18.51).

**Table 1:** Characteristics of the study participants

| Characteristics | Male (n = 70) |
| --- | --- |
| Demography | Mean ± SD |
| Age (years) | 51.34 ± 9.33 |
| Height (cm) | 170.38 ± 5.09 |
| Weight (kg) | 71.30 ± 10.10 |
| BMI (kg/m<sup>2</sup>) | 24.47 ± 3.00 |
| BSA (m<sup>2</sup>) | 1.83 ± 0.14 |
| HS |  |
| uHS (kg) | 29.80 ± 9.28 |
| uHS<sub>BMI</sub> | 1.24 ± 0.41 |
| uHS<sub>BSA</sub> | 16.31 ± 5.03 |
| Cardiorespiratory fitness |  |
| AT (mL/kg·min) | 10.29 ± 2.72 |
| VO<sub>2</sub>peak (mL/kg·min) | 16.12 ± 3.69 |
| HRrest (rpm) | 82.45 ± 11.64 |
| HRAT (rpm) | 103.75 ± 12.56 |
| HRmax (rpm) | 129.69 ± 19.78 |
| RERmax | 1.19 ± 0.14 |
| VE<sub>max</sub> (mL/min) | 46.99 ± 13.35 |
| Loadmax (W) | 71.90 ± 27.02 |
| Psysmax (mmHg) | 170.66 ± 21.84 |
| Pdiamax (mmHg) | 91.63 ± 13.71 |
| METmax | 4.61 ± 1.05 |
| VE/VCO<sub>2</sub> | 30.48 ± 5.09 |
| OUES | 1411.73 ± 295.94 |
| Type of stroke*N (%) |  |
| Hemorrhagic | 29 (40.85%) |
| Ischemic | 42 (59.15%) |
| Stroke location*N (%) |  |
| Left | 37 (52.11%) |
| Right | 34 (47.89%) |
| Hypertension*N (%) | 53 (74.65%) |
| Diabetes mellitus*N (%) | 24 (33.80%) |
| Barthel Index | 70.77 ± 18.51 |
| NIHSS | 3.32 ± 2.82 |

SD: standard deviation; BMI: body mass index; BSA: body surface area; uHS: handgrip strength on the unaffected side; uHSBMI: uHS adjusted for BMI; uHSBSA: uHS adjusted for BSA; AT: anaerobic threshold; VO<sub>2</sub>peak: peak oxygen uptake; HRrest: resting heart rate; HRAT: heart rate at anaerobic threshold; HRmax: max heart rate; loadmax: max work-load; RERmax: respiratory exchange ratio at max work-load; VEmax: minute ventilation at max work-load; Psysmax: systolic pressure at max work-load; Pdiamax: diastolic pressure at max work-load; METmax: metabolic equivalent at max workload; VE/VCO<sub>2</sub>: the minute ventilation/carbon dioxide production slope; OUES: oxygen uptake efficiency slope; BI: barthel index; NIHSS: national institutes of health stroke scale. Values are shown as mean ± SD or as number (%).

### 3.2. Univariate Correlations among Characteristics with VO_2_peak

Table 2 shows the association between the VO_2_peak and calculated variables. The most significant correlations (in descending order of significance) were observed between VO_2_peak by uHS_BSA_ (*p* < 0.001), uHS_BMI_ (*p* < 0.001), uHS (*p* = 0.008), BSA (*p* = 0.0211), and BMI (*p* = 0.0399). For strength, positive relationships were ascertained between VO_2_peak and uHS_BSA_ (R^2^=0.213, *p* < 0.001; Fig. 1a) as well as between VO_2_peak and uHS_BMI_ (R^2^=0.212, *p* < 0.001; Fig. 1b). In addition, NIHSS and BI were significantly correlated with the VO_2_peak (*p* < 0.01). No significant correlations were observed between the VO_2_peak and age, type of stroke, stroke location, hypertension, diabetes mellitus, and behavioral lifestyle of smoking and drinking. These results indicated that VO_2_peak was associated with the post-stroke function and HS on the unaffected side.

**Table 2:** Univariate correlations between participant characteristics and VO_2_peak

| Characteristics | VO<sub>2</sub>peak |  |
| --- | --- | --- |
|  | B (95% CI) | p-value |
| Age (years) | -0.075 (-0.167, 0.017) | 0.1128 |
| BMI | -0.301 (-0.583, -0.019) | 0.0399 |
| BSA | -6.997 (-12.807, -1.188) | 0.0211 |
| uHS | 0.155 (0.068, 0.241) | 0.0008 |
| uHSBMI | 4.111 (2.241, 5.980) | <0.0001 |
| uHSBSA | 0.339 (0.186, 0.493) | <0.0001 |
| Type of stroke |  |  |
| Hemorrhagic | Ref |  |
| Ischemic | 0.513 (-1.244, 2.270) | 0.5692 |
| Stroke location |  |  |
| Left | Ref |  |
| Right | -1.344 (-3.048, 0.359) | 0.1266 |
| NIHSS | -0.499 (-0.785, -0.213) | 0.0011 |
| BI | 0.105 (0.065, 0.145) | <0.0001 |
| Hypertension |  |  |
| None | Ref |  |
| Yes | -1.918 (-3.855, 0.020) | 0.0565 |
| Diabetes mellitus |  |  |
| None | Ref |  |
| Yes | -0.961 (-2.777, 0.854) | 0.3030 |
| Cardiorespiratory fitness |  |  |
| AT (mL/kg·min) | 0.961 (0.734, 1.188) | <0.0001 |
| HRrest (rpm) | 0.022 (-0.053, 0.096) | 0.5712 |
| HRAT (rpm) | 0.100 (0.035, 0.165) | 0.0037 |
| HRmax (rpm) | 0.102 (0.065, 0.139) | <0.0001 |
| RERmax | 6.719 (0.636, 12.803) | 0.0339 |
| VEmax (mL/min) | 0.197 (0.152, 0.243) | <0.0001 |
| Loadmax (W) | 0.040 (0.011, 0.070) | 0.0094 |
| Psysmax (mmHg) | 0.031 (-0.008, 0.070) | 0.1283 |
| Pdiamax (mmHg) | -0.025 (-0.088, 0.038) | 0.4427 |
| METmax | 3.513 (3.488, 3.539) | <0.0001 |
| VE/VCO<sub>2</sub> | 0.143 (-0.027, 0.313) | 0.1045 |
| OUES | 0.008 (0.006, 0.011) | <0.0001 |

BMI: body mass index; BSA: body surface area; uHS: handgrip strength on the unaffected side; uHSBMI: uHS adjusted for BMI; uHSBSA: uHS adjusted for BSA; AT: anaerobic threshold; VO<sub>2</sub>peak: peak oxygen uptake; BI: Barthel index; NIHSS: national institutes of health stroke scale; HRrest: resting heart rate; HRAT: heart rate at anaerobic threshold; HRmax: max heart rate; Loadmax: max work-load; RERmax: respiratory exchange ratio at max workload; VEmax: minute ventilation at max work-load; Psysmax: systolic pressure at max workload; Pdiamax: diastolic pressure at max work-load; METmax: metabolic equivalent at max workload; VE/VCO<sub>2</sub>: the minute ventilation/carbon dioxide production slope; OUES: oxygen uptake efficiency slope; BI: Barthel index; NIHSS: national institutes of health stroke scale; Scr: serum creatinine; UA: uric acid; CI: confidence interval; Ref: Reference.

**Figure 1:**
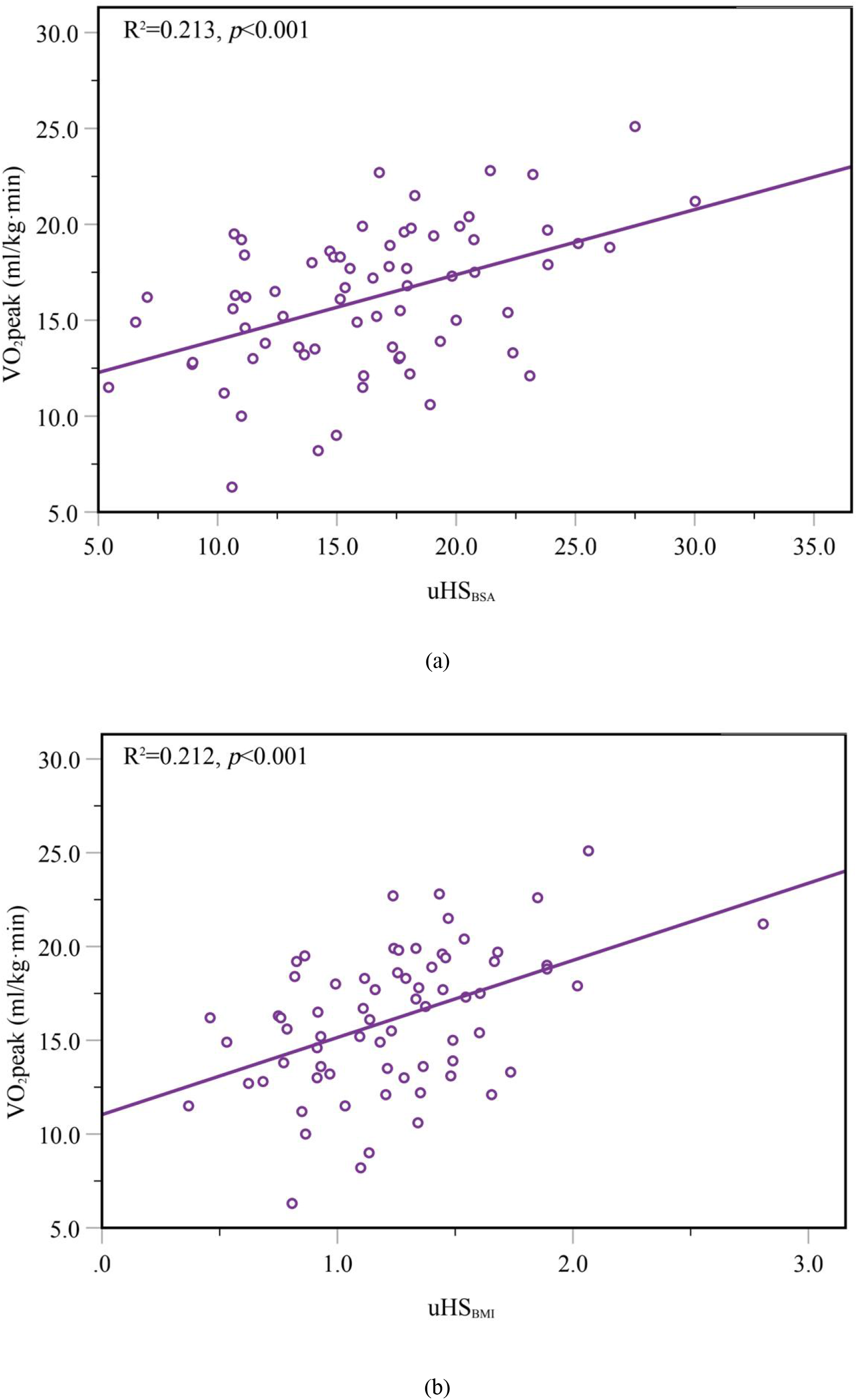
(a) Linear regression between uHS_BSA_ and the VO_2_peak in male patients (R^2^=0.213, *p*<0.001). (b) Linear regression between uHS_BMI_ and the VO_2_peak in male patients (R^2^=0.212, *p*<0.001).

### 3.3. Multiple Regression Analysis of Characteristics with VO_2_peak

A multiple linear regression analysis for VO_2_peak was applied (Table 3). The BI, uHS_BMI_, uHS_BSA_, and AT were significant factors for the independent prediction of the VO_2_peak. Age and physiological tests were included in the multivariate analysis to improve the proportion of variance explained by the model. The combination of age, BI, BSA, and uHS_BMI_ accounted for the greatest variance (R^2^ = 0.419). The addition of AT improved the predictive value and was included in the final multivariate model (R^2^ = 0.655). Figure 2 shows a plot of VO_2_peak calculated from the final multivariate model and VO_2_peak observed from exercise testing. A strong intraclass correlation [95% confidence interval (95% CI)] was observed between the actual and calculated VO_2_peak of 0.794 (95% CI: 0.687-0.867, *p* < 0.001, Figure 2). The calculated VO_2_peak estimated by the currently developed regression model did not differ systematically from the actual VO_2_peak (16.08 ± 2.98 vs. 16.13 ± 3.69 mL/kg/min, *p* = 0.841). According to the Bland-Altman analysis, the measured and calculated VO_2_peak were within good agreement, with 97.1% of the data points falling within the limits of agreement (−4.20–4.30) (Figure 3). The calculated VO_2_peak was marginally higher when the VO_2_peak was low. The difference is decreased as the CRF increased. The calculated VO_2_peak tends to lead to an underestimation of the VO_2_peak when the VO_2_peak is high. With a mean difference of 0.05 mL/kg/min, the calculated VO_2_peak was higher than the VO_2_peak by direct measurement. There was a mean absolute calculation error of 12.4% (95% CI: 10.12–14.62).

**Table 3:** Multivariate regression analysis on the association between participant characteristics and VO_2_peak

| Variable | Model 1 | Model 2 | Model 3 | Model 4 | Model 5 |
| --- | --- | --- | --- | --- | --- |
| Age | -0.044 | -0.025 | -0.039 | 0.009 | 0.003 |
| BI | 0.102† | 0.082† | 0.085† | 0.052† | 0.050† |
| BMI | - | -0.187 | - | - | - |
| BSA | - | - | -5.791* | - | - |
| uHSBMI | - | - | 2.232* | 2.290† | - |
| uHSBSA | - | 0.207† | - | - | 0.191† |
| AT(ml/kg·min) | - | - | - | 0.774† | 0.776† |
| Constant | 11.210 | 12.772 | 19.972 | 1.257 | 1.295 |
| Model adjusted R<sup>2</sup> | 0.268 | 0.359 | 0.383 | 0.631 | 0.634 |
| Model R<sup>2</sup> | 0.289 | 0.396 | 0.419 | 0.652 | 0.655 |

\*P < 0.05;
†.01.
R<sup>2</sup> = coefficient of determination.
uHS: handgrip strength on the unaffected side; BMI: body mass index; BSA: body surface area;
uHSBMI: uHS adjusted for BMI; uHSBSA: uHS adjusted for BSA; VO<sub>2</sub>peak: peak oxygen uptake; BI:
barthel index; NIHSS: national institutes of health stroke scale.

**Figure 2:**
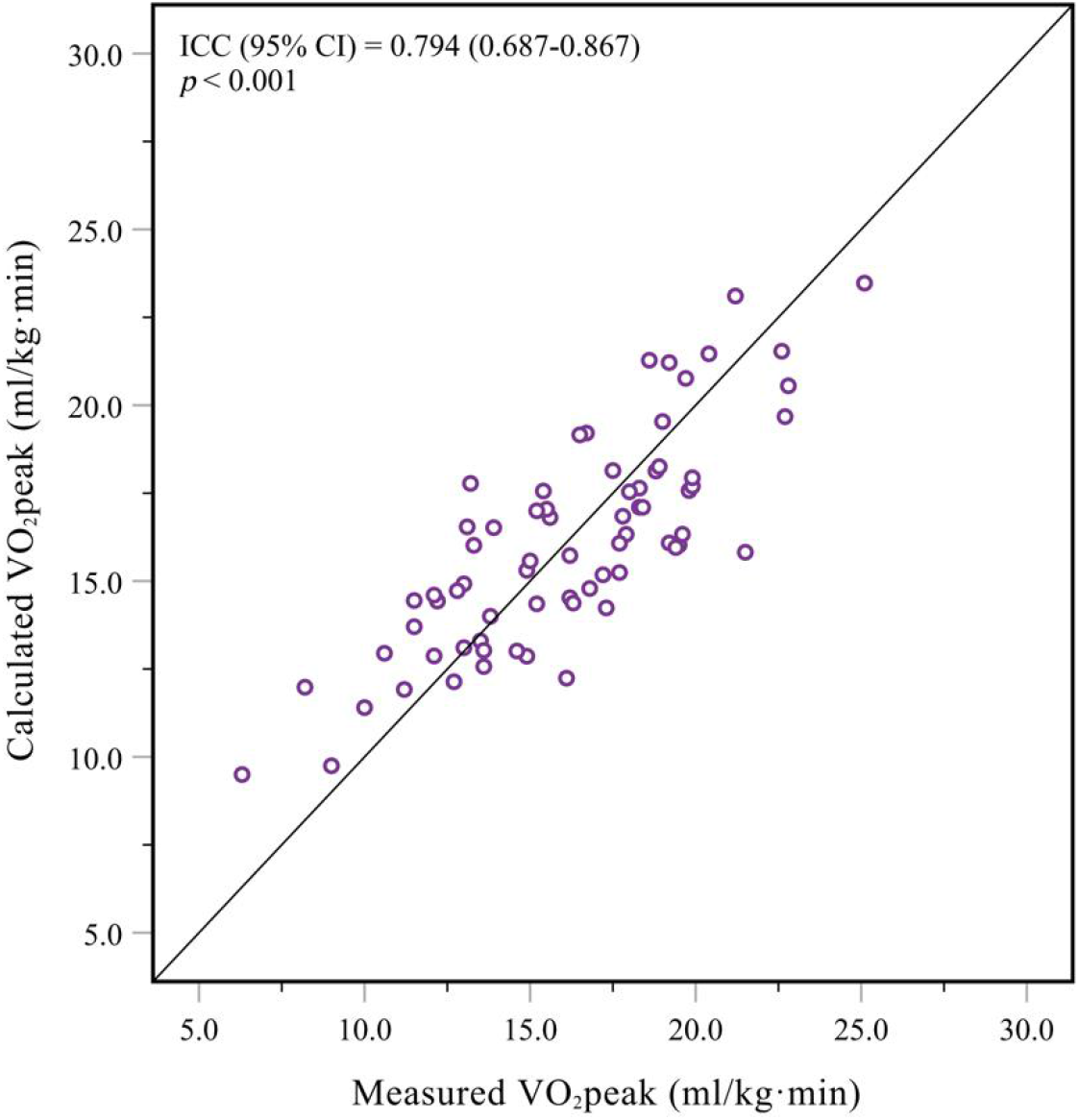
Plot of measured peak oxygen utilization (VO_2_peak; mL/kg·min) according to model-calculated VO_2_peak (mL/kg·min) estimated using the final multivariate model (Table 3, Model 5). The diagonal line represents the line of perfect agreement.

**Figure 3:**
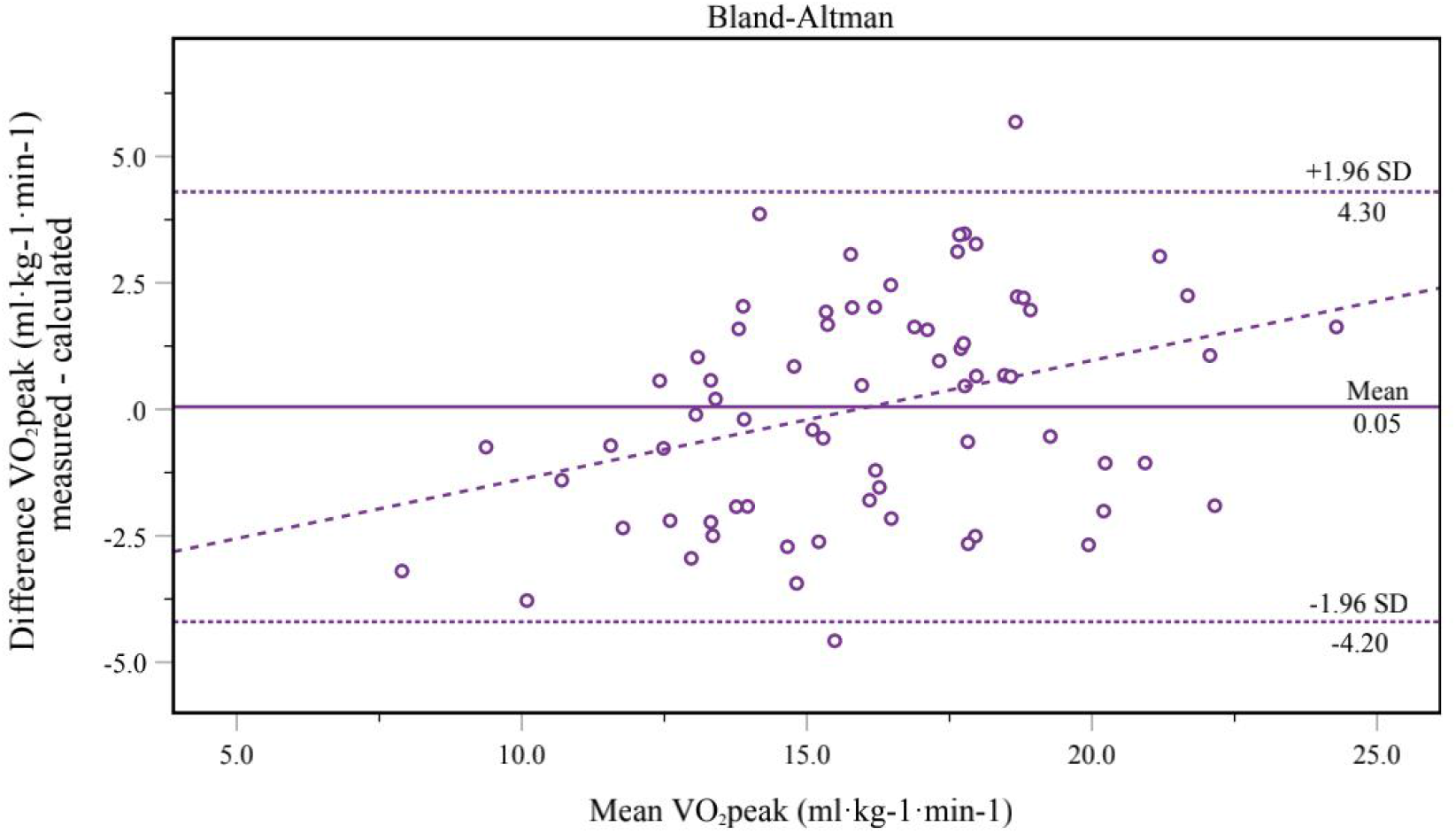
Bland-Altman plots showing agreement between the measured and calculated VO_2_peak by the final multivariate model (Table 3, Model 5). The solid and dotted lines indicate the mean difference and 95% limits of agreement (±1.96 × S.D. difference), respectively.

## Discussion

This study serves as an initial examination of the clinical and physiological determinants of cardiorespiratory fitness in patients with stroke. BI was found to be an independent predictor in multiple linear regression models for VO_2_peak^[21]^. The study findings also demonstrated that uHS_BMI_ and uHS_BSA_ are independent indicators of VO_2_peak in post-stroke male patients, even after adjustment for other significant covariant factors.^[8]^ Furthermore, the estimation of the VO_2_peak using linear regression, including the age, BI, BMI/BSA, and uHS_BSA_/uHS_BMI_, explained 35% or 38% of the variance in VO_2_peak, respectively. The addition of AT improved the predictive value of the model (R^2^ = 0.63). The results of this analysis suggest that a lower uHS is associated with lower levels of CRF in stroke patients, and grip strength measurement may be a practical risk screening tool in this population of patients.

Evidence has confirmed the relationship between CRF and incident hypertension and diabetes mellitus in the past few years^[22]^. However, individuals with a history of hypertension or diabetes mellitus did not have a significantly low VO_2_peak^[23]^. It is well-established that CRF typically declines after a stroke.^[6]^ The findings of the current study demonstrate that patients with stroke exhibit peak oxygen uptake (VO_2_peak) that is less than 60% of that in a healthy, age- and gender-matched population. Additionally, no clear relationship was observed between the VO_2_peak and age. In our study, we recruited participants with mild stroke, with the NIHSS mean score being 3.32 ± 2.82. Possibly, cardiorespiratory fitness is impaired even more severely after a severe stroke. The maximal oxygen uptake may have been overestimated, as less healthy and older stroke survivors may not be tolerant to maximal exercise testing.

According to these findings, BI is significantly associated with cardiovascular fitness in male patients of stroke. The BI is a reliable and valid scale for measuring ADL in patients of stroke.^[24]^ A previous study demonstrated that the baseline cardiovascular fitness correlates significantly with ADL function and can serve as an important prognostic factor for functional recovery after subacute stroke.^[21]^ Baert et al. also observed a significant correlation between the VO_2_peak and BI (r = 0.47 and 0.59, respectively) at 6 and 12 months poststroke.^[7]^ This is probably because upper-level ADLs require higher aerobic capacity for completion.^[25]^ In addition, disability after stroke usually increases the energy requirement of all activities based on gross motor inefficiencies.

In addition to measures of ADL, the anaerobic threshold was significantly correlated with CRF post stroke. VO2peak, the highest oxygen uptake obtained during exercise, may be substantially influenced by motor function after stroke because graded exercise tests (GXTs) can be restricted by motor dysfunction (e.g., paresis).^[26]^ Unlike VO2peak, AT is a parameter of the submaximal oxygen uptake that can be safely assessed at submaximal exercise levels and correlated with endurance performance.^[27,28]^ AT has been suggested to be a reliable index of aerobic capacity since it assesses the capacity to perform sustained aerobic activity.^[29]^ In a previous study of 98 individuals poststroke, only 18% achieved VO_2max_, whereas 67% achieved AT.^[30]^ Although the anaerobic threshold is easier to attain than the peak oxygen uptake, the peak oxygen uptake is irreplaceable as a universal prognostic marker.^[31]^

Of note, uHS_BMI_/uHS_BSA_ was an independent indicator of CRF after stroke, whereas uHS_BMI_/uHS_BSA_ has never been previously reported to play this role in patients of stroke. Several studies have shown the relationship between HS and VO_2_peak, not only in patients with congestive heart failure (CHF) and chronic obstructive pulmonary disease (COPD) but also in human immunodeficiency virus (HIV)-infected patients and paraplegic men (respectively, r^2^ =0.32, r =0.743, r = 0.41, r = 0.674, all, p < 0.01).^[13–15,32]^ In addition, we observed positive correlations between HS_BMI_/HS_BSA_ and VO_2max_ in young adults (p < 0.05), which was in agreement with the findings of the current study.^[8]^ Although the relative values of uHS adjusted for BMI and BSA showed a stronger correlation with VO_2peak_ than the absolute values in this study, the correlation in patients with stroke was lower than that in other chronically ill patient populations, which may be due to the impairment of the motor function of the unaffected limbs of patients after stroke.^[33]^ HS is a simple and non-invasive measurement of overall muscle strength as well as function. Although no study has reported the mechanism of association between CRF and HS, evidence from early investigations suggested that the decline in VO_2peak_ may be attributed to a decrease in the peripheral O_2_-diffusive capacity in skeletal muscles. One major factor that affects the VO_2_peak is the cardiac output, which increases during handgrip. This could explain the association between HS and CRF.

Meanwhile, these findings showed that the predictive value of the final multivariate model (R2 = 0.656) was similar to that in HIV-infected patients (R2 = 0.63) but lower than that in paraplegic men (R2 = 0.811).^[15]^ Peniche et al. showed that the equations using age, sex, physical activity, test mode, and anthropometric variables cannot show adequate validity to predict the VO_2_max in individuals after chronic stroke. The results of the present study can help identify the physiological and clinical factors that influence cardiorespiratory fitness. Moreover, the clinical and physiological parameters in the current research model can be referenced for patients with stroke when clinicians are unable to use gold-standard methods to assess CRF.

A limitation of this study was the characteristics of the study population. Because findings from our previous study have indicated a weak association between HS_BMI_/HS_BSA_ and CRF in women and they were not well represented in patients with stroke, they were not included in the analysis. Patients with bilateral hemisphere stroke and severe functional limitations were excluded from the study.^[30]^ Therefore, the current study was limited to male patients without severe stroke, and the sample size was relatively small; any conclusions outside this population should be made with caution. Conversely, we did not include people with major cognitive impairments or difficulties to communicate. Finally, because this was a cross-sectional study, causation cannot be determined, but can be verified by prospective studies in the future.

## Conclusion

Our findings demonstrate that uHS_BMI_ and uHS_BSA_ are the predictors of cardiorespiratory fitness in male patients with stroke. The multivariate regression model comprising AT, handgrip strength, and anthropometric variables showed a strong correlation with the VO_2_peak. Since AT could be relatively easily obtained at the submaximal exercise testing measure, we suggest that uHS_BMI_/uHS_BSA_ combined with AT and anthropometric variables could serve as indicators of VO_2_peak. HS on the unaffected side offers a safe, feasible, and inexpensive measure of fitness in male patients with stroke, but a larger and more diverse study in a stroke-patient population is necessary for validating its use as a prognostic indicator.

## Data Availability

Data is available on request from the authors.

## Acknowledgement

We thank the staff from our institution who actively participated in the current study.

